# Comparison of phylogenetic metrics of transmission in symptomatic and asymptomatic tuberculosis

**DOI:** 10.1101/2025.08.27.25334397

**Authors:** Kesia Esther Da Silva, Paulo César Pereira dos Santos, Daniel Henrique Tsuha, Katharine S. Walter, Eunice Atsuko Totumi Cunha, Caroline Colijn, Ted Cohen, Roberto Dias de Oliveira, José Victor Bortolotto Bampi, Mariana G Croda, Crhistinne Cavalheiro Maymone Gonçalves, Luiz Henrique Ferraz Demarchi, Julio Croda, Jason R. Andrews

## Abstract

**Background:** Understanding drivers of *Mycobacterium tuberculosis* (*Mtb*) transmission remains a critical challenge in high-burden settings. Tuberculosis control efforts traditionally target symptomatic individuals, yet the role of asymptomatic cases in sustaining transmission is increasing recognized.

**Methods:** We conducted a genomic and epidemiological analysis of *Mtb* isolates collected in Mato Grosso do Sul, Brazil, between 2008 and 2024. From 2017 to 2022, active case finding was performed in three of the state’s largest prisons, whereby sputum was collected from individuals irrespective of symptoms and tested by GeneXpert and culture. We evaluated several metrics of recent transmission from symptomatic and asymptomatic individuals, including phylogenetic clustering, Time-scaled Haplotype Density (THD), Local Branching Index (LBI), and transmission probabilities inferred using the Bayesian Reconstruction and Evolutionary Analysis of Transmission Histories (BREATH).

**Findings:** We sequenced 2,362 *Mtb* strains, of which 3.5% (115/2,362) were resistant to at least one drug, and 0.6% (16/2,362) were multi-drug resistant. Most strains were lineage 4, and 78.2% of all isolates were part of a genomic cluster. Among 2,362 individuals with tuberculosis, 1,137 were incarcerated at the time of diagnosis. Among these, 505 were identified through active case finding: 277 had symptomatic disease and 228 had asymptomatic tuberculosis. There was no significant difference in phylogenetic clustering proportion (77% vs. 85%; p= 0.816), THD (median 0.50 vs. 0.39; p = 0.120), or LBI (median 0.00863 vs. 0.00871; p = 0.086) between symptomatic and asymptomatic individuals. Bayesian transmission trees revealed no significant difference in the number of secondary infections inferred from symptomatic compared with asymptomatic individuals (p = 0.56). These findings were consistent across genomic clusters and robust to model assumptions.

**Interpretation:** We identified no differences in transmission from symptomatic compared with asymptomatic individuals, using several genomic measures of transmission, underscoring the substantial contribution that asymptomatic tuberculosis makes to transmission at the population level.

**Evidence before this study:** We searched PubMed from inception to June 1, 2025, without language restrictions, using the terms “tuberculosis”, “asymptomatic”, “transmission”, “infectiousness” and “genomic epidemiology”. We also reviewed reference lists of relevant studies and reports from the WHO Global Tuberculosis Programme. Most available evidence on the contribution of asymptomatic tuberculosis to transmission comes from cross-sectional contact studies, which typically use tuberculin skin tests or interferon gamma release assays to measure *Mycobacterium tuberculosis* infection risk in contacts. These studies have generally found no major differences in infection risk between contacts of symptomatic and asymptomatic individuals, but they measure lifetime infection risk and cannot establish the source of exposure. Few studies have used genomic epidemiology to directly assess transmission by symptom status, and those that exist have been small in scale and limited in scope. Mathematical modelling has suggested that asymptomatic individuals could account for a substantial proportion of transmission, but empirical, population-level data from high-incidence settings remain scarce.

**Added value of this study:** We combined genomic, epidemiological, and clinical data from over 2,362 *M. tuberculosis* isolates collected in Mato Grosso do Sul, Brazil, including more than 500 cases identified through active case finding in prisons, to directly compare multiple genomic metrics of transmission between symptomatic and asymptomatic individuals. We found no significant difference in genomic clustering, phylogenetic epidemic success, or the number of estimated secondary infections between groups. Our study is among the largest to date to evaluate transmission resulting in tuberculosis disease by symptom status. These findings provide robust, population-based evidence that asymptomatic tuberculosis can contribute to transmission at levels comparable to symptomatic disease, even in settings with extensive case finding.

**Implications of all the available evidence:** Our findings, in combination with previous evidence, indicate that symptom-based case detection strategies are insufficient to substantially reduce tuberculosis transmission. In high-burden settings, systematic screening irrespective of symptoms, is essential to identify and treat infectious cases earlier. Public health programmes should prioritize expanding active case finding in both high-risk institutional settings and the community to capture asymptomatic individuals who may sustain transmission.

## Introduction

Tuberculosis (TB) remains the leading cause of infectious disease mortality worldwide, responsible for an estimated 1.25 million deaths annually. Despite global commitment to the End TB Strategy, progress has been slow, with only a 2% annual decline in incidence (1). One of the main obstacles to reducing the burden of tuberculosis is the continuous generation of new infections, primarily by transmission from people with undiagnosed disease. About half of individuals with tuberculosis at any given time lack symptoms, and it is increasingly recognized that such individuals may play an important role in transmission.

However, the relative contribution of asymptomatic and symptomatic individuals to *Mycobacterium tuberculosis* (*Mtb*) transmission at the population level remains unclear. Studies evaluating risk of *Mtb* infection, as measured by tuberculin skin tests or interferon gamma release assays, among close contacts have found no significant differences in infection risk between individuals exposed to symptomatic versus asymptomatic individuals with tuberculosis (2,3). However, these studies, while providing valuable data, have some important limitations. As cross-sectional studies with an outcome that is a binary measure of *Mtb* infection reflecting cumulative life exposure, it is not clear whether the *Mtb* infections in contacts were due to transmission from the putative index case as opposed to other lifetime exposures, including potential shared exposures. Further, not all exposures result in equal risk of progression to tuberculosis disease; understanding the contribution of asymptomatic tuberculosis in generating cases of tuberculosis disease would provide more direct evidence for its role in the sustaining the epidemic.

Mathematical models demonstrate that understanding the relative infectiousness of asymptomatic tuberculosis is critical to forecasting the impact of case finding strategies and, accordingly, to prioritizing allocation of diagnostic resources most effectively (4). To address this knowledge gap, we evaluated genomic evidence of transmission from individuals with tuberculosis identified through active case finding—conducted irrespective of symptoms—in high tuberculosis burden prisons, comparing multiple metrics of transmission in individuals with symptomatic and asymptomatic tuberculosis.

## Methods

### Study population

We conducted population-based genomic surveillance for tuberculosis in the state of Mato Grosso do Sul in Central-West Brazil, from 2008 through 2024. Surveillance efforts included both active and passive case finding, focusing on the state’s two largest cities, Campo Grande and Dourados. Active case finding was carried out in three largest prisons of the state from 2017 to 2024. Incarcerated individuals were screened regardless of symptoms, and screening procedures included a symptom assessment, chest radiography, and sputum testing using Xpert MTB/RIF and culture for positive Xpert MTB/RIF. Sputum was collected from all individuals able to produce a sample (S1 Text).

A TB case was defined by a positive sputum test by Xpert® MTB/RIF Ultra. Cases were further classified as symptomatic or asymptomatic TB based on responses to standardized symptom questions administered by a team of trained nurses during screening. Symptomatic TB was diagnosed in individuals presenting with one or more of the specified TB symptoms, irrespective of their duration, while asymptomatic TB referred to cases without any of these TB symptoms. TB-related symptoms were defined as cough of any duration, sputum, fever, appetite loss, night sweats, chest pain, trouble breathing, and weight loss of any duration.

We included metadata and sequencing data from all participants to generate phylogenetic trees and for all analyses of clustering, transmission metrics, and transmission tree inference. However, for the comparisons of symptomatic and asymptomatic tuberculosis, we restricted the analysis to incarcerated individuals, because very few individuals diagnosed in the community had asymptomatic tuberculosis (active case finding is not routinely performed). Restricting to incarcerated individuals identified through symptom-agnostic active case finding mitigates the effects of symptoms on diagnosis.

### Whole genome sequencing and bioinformatics analyses

Whole-genome sequencing was performed on the Illumina NovaSeq X Plus platform (2 × 150 bp). We analyzed genomic variation using a publicly available pipeline available on GitHub (https://github.com/ksw9/mtb-call2?) (S1 Text). Lineages and evidence of mixed infection were inferred using TBProfiler v4.2.0, which identifies over 1,000 lineage-defining SNPs. TBProfiler was also used to detect mutations associated with drug resistance (5).

### Phylogenetic reconstruction

We constructed full-length consensus FASTA sequences from VCF files and used SNP-sites to extract a multiple alignment of internal variant sites only (6). To construct the phylogeny, we identified the best-fit substitution model using ModelFinder (7) (S1 Text). A maximum likelihood (ML) phylogenetic tree was then inferred using IQ-TREE, with 1,000 ultrafast bootstrap replicates to assess node support. We used the R package *ape* (8) to measure pairwise differences between samples and genomic clusters were defined using a commonly applied threshold of 12 SNPs for pairwise distance (9). We compared the proportion of isolates clustering from symptomatic versus asymptomatic participants using Chi-squared test.

### Phylogenetic metrics of transmission

We used the time-scaled haplotypic density (THD) method to evaluate the influence of clinical symptoms and incarceration status on the epidemic success of *M. tuberculosis* isolates. THD computes an independent success index for each isolate in a collection, enabling the identification of predictors of success (10). THD estimates were calculated from pairwise SNP distance matrices using a timescale of 20 years, an effective genome size of 4 × 10⁶ bp, and an average per-nucleotide substitution rate of 5.8 × 10⁻⁸ (11). We used generalized linear models to examine the influence of symptoms and incarceration status on the THD success index.

Next, we estimated the Local Branching Index (LBI) (12) for each tip in the maximum ML phylogeny using the R package TreeImbalance (13). LBI is a quantitative measure of pathogen fitness inferred from the shape of the phylogenetic tree. It is based on the total branch length surrounding each node (both internal and terminal), with contributions discounted exponentially as distance from the node increases. Higher LBI values indicate more rapid expansion and are associated with increased fitness (14). We computed LBI values for all nodes in the ML tree using this scaling factor. We compared THD and LBI distributions from symptomatic versus asymptomatic individuals using a Wilcoxon rank sum test.

### Transmission inference

To investigate transmission patterns in the ten largest genomic clusters, we first grouped *M. tuberculosis* isolates using a 12-SNP threshold. For each cluster, we constructed time-calibrated phylogenetic trees using BEAST v2.7.6 (15), incorporating sample collection dates to calibrate the tree tips. While phylogenies capture evolutionary relatedness between sampled isolates, they do not directly represent transmission events, especially in the presence of incomplete sampling and within-host diversity. To infer likely transmission events, including those involving unsampled hosts, we used the Bayesian Reconstruction and Evolutionary Analysis of Transmission Histories (BREATH) model (16). BREATH allows simultaneous inference of both phylogeny and transmission history, incorporating epidemiological data. We configured the analysis in BEAUti, setting the tree prior to BREATH and the site model to HKY+Γ4+I. After sensitivity analysis (**S1 Text**), the sampling hazard parameters were set as shape = 10, rate = 6.5, and C = 0.75. For the transmission hazard, we set shape = 10, rate = 8.5, and C = 2. The molecular strict clock was bounded between 4.1 × 10⁻⁸ and 1.33 × 10⁻⁷ substitutions/site/year and a log-normal prior on the substitution rate (mean = −17, SD = 2) consistent with prior estimates for *M. tuberculosis* Lineage 4 (17).

From the inferred transmission trees, we used the WIWVisualiser app to generate visualizations of who-infected-whom relationships. We also generated a pairwise transmission probability matrix summarizing the likelihood of direct transmission between every pair of individuals. Using this matrix, we calculated the probability-weighted number of secondary infections attributable to each case. We then compared these distributions between symptomatic and asymptomatic individuals to assess differences in transmission potential using a Wilcoxon rank sum test. Statistical analyses were performed to evaluate the association between clinical symptoms and transmission, adjusting for relevant covariates.

### Role of the funding source

The funder of the study had no role in study design, data collection, data analysis, data interpretation, or writing of the report.

## Results

### Genomic surveillance of *M. tuberculosis* in Mato Grosso do Sul, Brazil

Between 2002 and 2024, *M. tuberculosis* isolates from 2,959 individuals residing in Mato Grosso do Sul were sequenced (Figure 1). After excluding 165 genomes with low sequencing coverage, 2,794 high-quality isolates remained. An additional 91 isolates were excluded due to contamination or evidence of mixed infection. Among the remaining genomes, 2,696 (99.7%) were classified as belonging to lineage 4. Given the predominance of this lineage, we focused our analysis on lineage 4 isolates. After further excluding samples with missing metadata, 2,362 lineage 4 isolates were included in the final analysis. Of these isolates, 1,137 (48.1%) were from individuals incarcerated at the time of TB notification, 116 (4.9%) were from individuals residing in the community who had previously been incarcerated, and 1,016 (43.0%) were from individuals in the community with no incarceration history; incarceration status was unknown for the remaining 93 (4.0%) individuals. From 2017 to 2024, 505 isolates were obtained from individuals through active case finding, with symptom data available; among these, 277 were from symptomatic participants and 228 were from asymptomatic participants.

**Figure 1.**
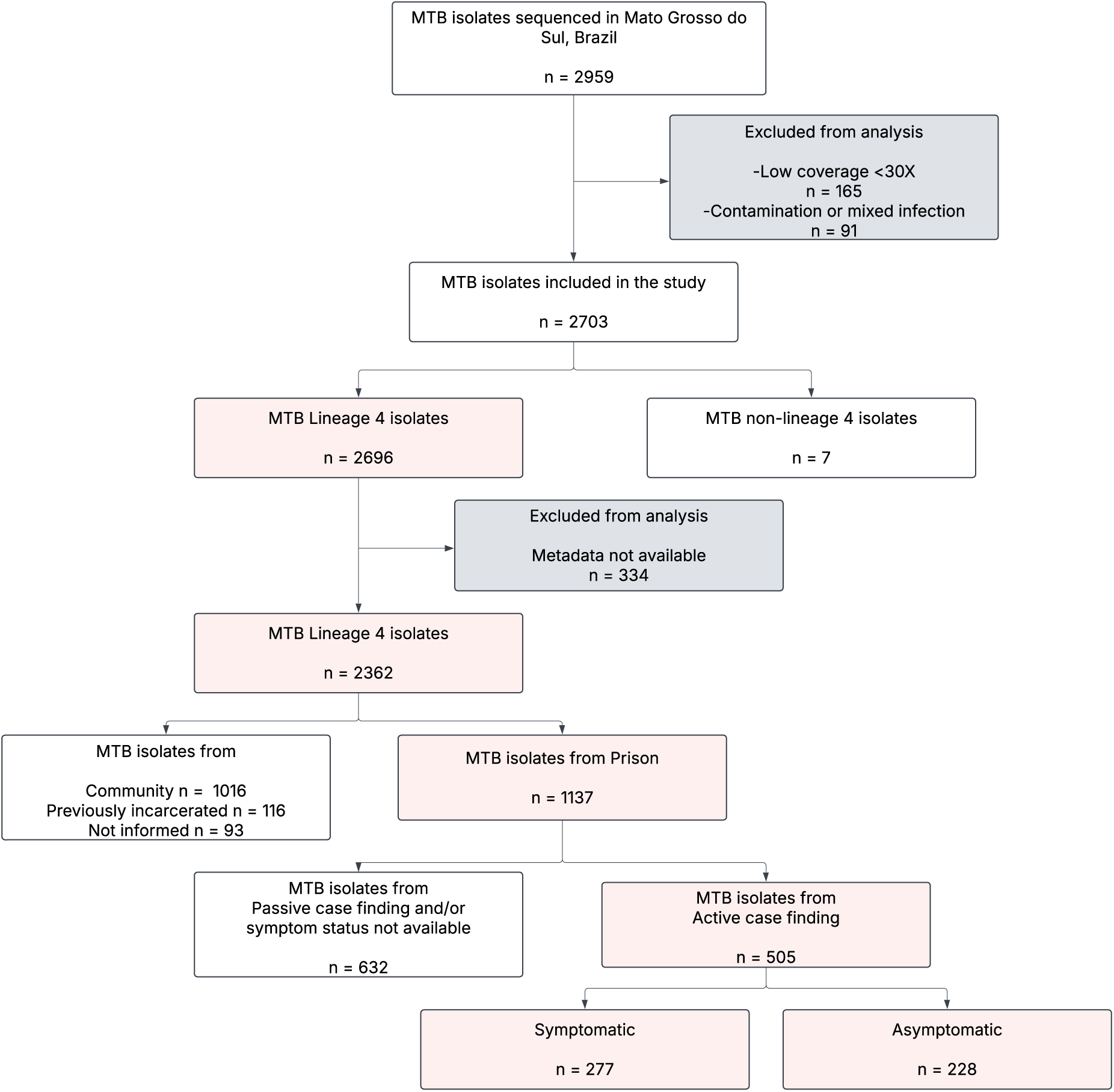
Flowchart describing genomic surveillance for *M. tuberculosis* in Mato Grosso do Sul, Brazil. Pink boxes indicate the subset of isolates included in the transmission analysis presented in this study.

### Phylogenetic structure and drug resistance of *M. tuberculosis*

Phylogenetic analysis of 2,362 *M. tuberculosis* lineage 4 isolates revealed a diverse population structure comprising multiple sub-lineages (Figure 2). The most prevalent sub-lineage was 4.1.2.1, accounting for 26.6% (628/2,362) of all isolates, followed by 4.3.3 (22.0%; 520/2,362), 4.4.1.1 (19.6%; 462/2,362), 4.3.4.2 (9.9%; 234/2,362), and 4.4.1.2 (8.0%; 188/2,362). The proportion of sequences collected from each lineage remained consistent throughout the study period (Figure S1). This distribution indicates the co-circulation of multiple sub-lineages with evidence of local clonal expansion, particularly among the dominant groups. The majority of isolates (95.1%, 2,247/2,362) were drug sensitive. Isoniazid monoresistance (HR-TB) was observed in 2.6% (61/2,363), rifampicin monoresistance (RR-TB) was found in 0.7% (71/2,363) and 0.6% (16/2,362) of isolates were classified as multidrug resistant (MDR-TB). An additional 0.9% (21/2,362) of isolates were classified as having other resistance patterns.

**Figure 2.**
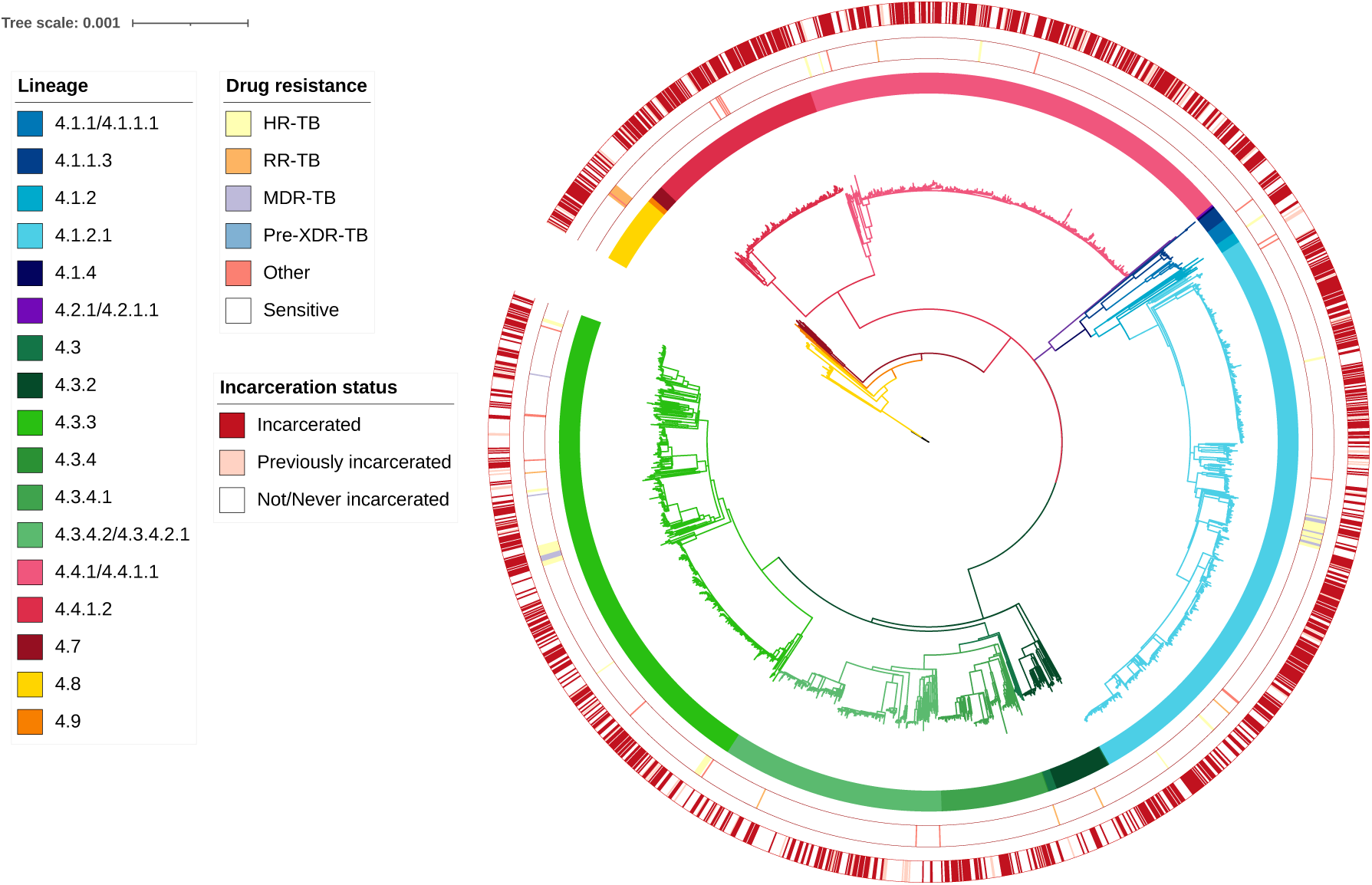
A maximum likelihood phylogeny of 2362 *M. tuberculosis* isolates from Lineage 4. Branch lengths are in units of substitutions per site. Branches are colored by sub-lineage. From the inside, rings are colored by sub-lineage, antimicrobial resistance prediction, and incarceration status at time of TB notification. “Other” resistance indicates resistance to at least one antibiotic other than isoniazid or rifampicin.

Isolates from incarcerated and non-incarcerated individuals were distributed throughout the phylogenetic tree and did not form distinct clades, suggesting recent and ongoing transmission between prison and community settings. *M. tuberculosis* diversity was largely shaped by the circulation of highly related clones. Cluster analysis using a 12-SNP threshold revealed that 78.2% (1,849/2,362) of isolates belonged to one of 198 genomic clusters. Clustering was higher among incarcerated individuals (83.8%) and those previously incarcerated (75.9%), compared to individuals in the community (71.1%) (Chi-square test, p < 0.0001).

### Characteristics of participants with symptomatic and asymptomatic tuberculosis

We analyzed clinical and demographic data from individuals diagnosed with TB, stratified by symptom status to assess factors associated with asymptomatic presentation (Table 1). There was no significant difference in age between asymptomatic and symptomatic individuals (median age: 29.5 [25–38] vs. 30 [26–35.7] years, p = 0.859). A greater proportion of asymptomatic individuals had less than eight years of schooling (87.3% vs. 72.6%, p < 0.001). Chest radiography findings were similar across both groups, with 87.1% of participants showing abnormal results (p = 0.807). Bacterial load measured by Xpert MTB/RIF differed significantly between groups (p < 0.001). Symptomatic individuals were more likely to have high (21.3%) or medium (27.4%) loads, whereas asymptomatic individuals were more likely to have low (47.4%) or very low (12.3%) loads. There were no significant differences in the prevalence of comorbidities such as HIV infection (p = 0155) or diabetes (p = 0.501). Previous incarceration (p = 0.295) was not significantly associated with symptom status. Behavioral risk factors were more prevalent in the asymptomatic group, including alcohol use (62.3% vs. 36.8%; p < 0.001), smoking (81.6% vs. 61.7%; p < 0.001), and drug use (75.0% vs. 64.3%; p = 0.019). Isoniazid resistance (HR-TB) was more frequently observed in asymptomatic individuals (4.4% vs. 0.7%; p = 0.001). Among symptomatic individuals, the most commonly reported symptom was cough, present in 89.9% of individuals, followed by chest pain (45.8%), fever (37.5%), weight loss (30.1%), difficulty breathing (30.3%), night sweats (27.1%), and loss of appetite (23.8%).

**Table 1.** Demographic and clinical characteristics of symptomatic and asymptomatic individuals with tuberculosis

| Characteristics | Total n (%) | Asymptomatic n (%) | Symptomatic n (%) | p value* |
| --- | --- | --- | --- | --- |
| **Age (median [IQR])** | 30 [25, 37] | 29.5 [25, 38] | 30 [26, 35.7] | 0.859 |
| **Ethnicity** |  |  |  | 0.022 |
| Mixed | 304 (60) | 141 (61.8) | 161 (58.1) |  |
| White | 90 (17.8) | 33 (14.5) | 57 (20.6) |  |
| Black | 56 (11.1) | 35 (15.4) | 21 (7.6) |  |
| Indigenous | 13 (2.6) | 5 (2.2) | 8 (2.9) |  |
| Asian | 6 (1.2) | 2 (0.9) | 4 (1.4) |  |
| **Schooling** |  |  |  | <0.001 |
| <8 years of schooling | 402 (79.3) | 199 (87.3) | 201 (72.6) |  |
| **Chest radiography** |  |  |  | 0.807 |
| Abnormal | 440 (87.1) | 201 (88.2) | 239 (86.3) |  |
| Normal | 9 (1.8) | 4 (1.8) | 5 (1.8) |  |
| **Bacterial load (Xpert)** |  |  |  | <0.001 |
| High | 78 (15.4) | 19 (8.3) | 59 (21.3) |  |
| Medium | 113 (22.3) | 37 (16.2) | 76 (27.4) |  |
| Low | 176 (34.7) | 108 (47.4) | 68 (24.5) |  |
| Very low | 44 (8.7) | 28 (12.3) | 16 (5.8) |  |
| Trace | 3 (0.6) | 2 (0.9) | 1 (0.4) |  |
| **Previously arrested** |  |  |  | 0.295 |
| Yes | 298 (59.0) | 130 (57.0) | 168 (60.6) |  |
| **Alcohol use** |  |  |  | <0.001 |
| Yes | 244 (48.3) | 142 (62.3) | 102 (36.8) |  |
| **Smoking** |  |  |  | <0.001 |
| Yes | 357 (70.7) | 186 (81.6) | 171 (61.7) |  |
| **Drug use** |  |  |  | 0.019 |
| Yes | 349 (69.1) | 171 (75.0) | 178 (64.3) |  |
| **Diabetes** |  |  |  | 0.501 |
| Yes | 14 (2.8) | 8 (3.5) | 6 (2.2) |  |
| **HIV status** |  |  |  | 0.155 |
| Positive | 11 (2.2) | 4 (1.8) | 7 (2.5) |  |
| **Drug resistance** |  |  |  | 0.001 |
| Isoniazid | 12 (2.4) | 10 (4.4) | 2 (0.7) |  |
| Rifampicin | 1 (0.2) | 0 (0.0) | 1 (0.4) |  |
| MDR-TB | 1 (0.2) | 0 (0.0) | 1 (0.4) |  |
| Other | 3 (0.6) | 1 (0.4) | 2 (0.7) |  |
| Sensitive | 488 (96.6) | 217 (95.2) | 271 (97.8) |  |

\* p-values generated using Chi-square test and t test.

### Determinants of recent *M. tuberculosis* transmission

Among individuals identified through active case finding, 80.7% (408/505) of isolates were part of 98 genomic clusters. Cluster proportions did not differ significantly between symptomatic (77%; 56 clusters) and asymptomatic (85%; 42 clusters) individuals (p = 0.372). After adjusting for sampling date, there remained no significant difference in clustering proportions between symptomatic and asymptomatic individuals (p = 0.742).

We evaluated the epidemic success of *M. tuberculosis* isolates using two phylogenetic metrics: time-scaled haplotype density (THD) and the Local Branching Index (LBI) (Figure 3). There was no significant difference in THD between isolates from asymptomatic (median 0.50, IQR 0.09-0.65) and symptomatic individuals (median 0.39, IQR 0.06-0.62) (p = 0.12). LBI estimates were slightly higher among asymptomatic individuals (median = 0.00863; range: 0.00248–0.0118) than among symptomatic individuals (median = 0.00871; range: 0.00224–0.0115), but this difference was not statistically significant (p = 0.085). To investigate predictors associated with epidemic success and increased fitness, we fit generalized linear models for both THD and LBI, adjusting for the date of TB notification and strain lineage. Incarceration status was a significant predictor of higher epidemic success across both metrics. For THD, isolates from incarcerated individuals had the highest values (β= 0.60, p < 0.001), followed by those from previously incarcerated individuals (β = 0.33, p = 0.006), compared to individuals with no incarceration history. A similar pattern was observed for LBI, with isolates from incarcerated individuals showing significantly higher estimates (β = 0.00071, p < 0.001). followed by previously incarcerated individuals (β = 0.00026, p = 0.15). Symptom status was not significantly associated with either THD (β = 0.01, p = 0.78) or LBI (β = 0.00006, p = 0.46).

**Figure 3.**
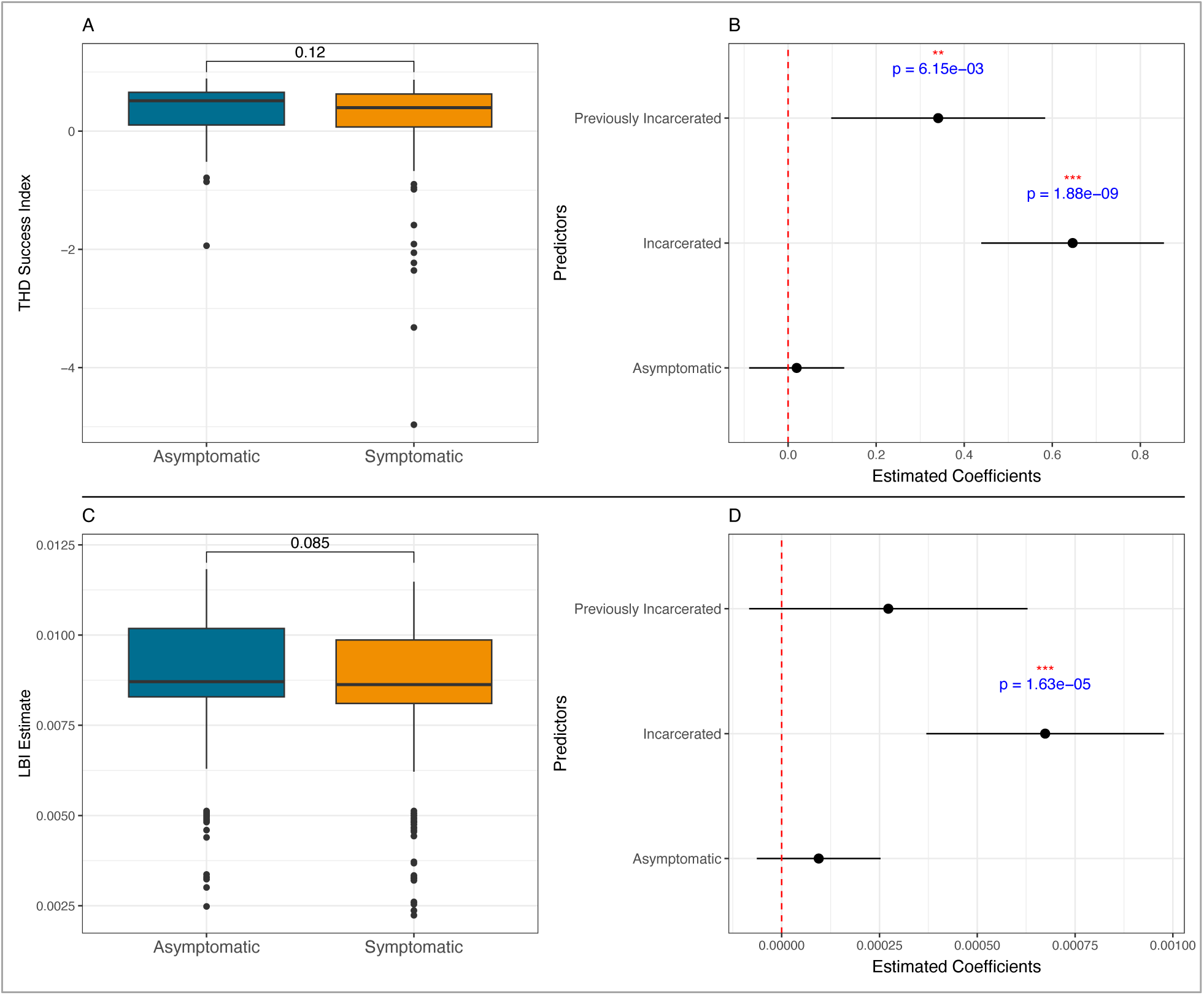
Metrics of epidemic success and pathogen expansion by symptom status. **(A)** Comparison of the Time-Scaled Haplotype Density (THD) Success Index between symptomatic and asymptomatic individuals. **(B)** Estimated coefficients from a Generalized Linear Model (GLM) evaluating the association between THD and predictors including symptom status and incarceration history. The reference group for previous incarceration and incarcerated was never incarcerated, and the reference group for asymptomatic was symptomatic. Error bars represent 95% confidence intervals, with significant predictors marked and p-values shown. **(C)** Comparison of Local Branching Index (LBI) estimates between symptomatic and asymptomatic individuals. **(D)** Estimated coefficients from a Generalized Linear Model (GLM) evaluating the association between LBI and predictors including symptom status and incarceration history. Error bars represent 95% confidence intervals, with significant predictors marked and p-values shown.

### Comparison of *M. tuberculosis* transmission from symptomatic versus asymptomatic individuals

We characterized transmission patterns across the ten largest genomic clusters using posterior distributions of transmission trees inferred with the BREATH model (16). This approach allowed inference of likely who-infected-whom relationships while accounting for unsampled hosts and timing of infections. To test the robustness of our findings, we compared the distributions of total transmission probabilities for symptomatic and asymptomatic individuals under two plausible scenarios that differ in how quickly transmission is assumed to occur relative to the speed of case detection. Estimated numbers of secondary infections attributed to symptomatic and asymptomatic individuals were similar across the two scenarios (Figure S2). Based on these patterns we selected the medium scenario (transmission mean = 1.5) for all downstream analyses, given this scenario reflects a more biologically and epidemiologically realistic assumption for *M. tuberculosis* in high-burden populations.

To quantify the contribution of asymptomatic individuals to transmission we estimated the average number of secondary infections attributable to each sampled individual from the inferred transmission trees. While this approach explicitly models unobserved cases, we calculated secondary infections among observed cases (sequenced isolates) due to higher confidence in transmission events. Among individuals with symptoms, the median number of estimated secondary infections was 0.37 (IQR = 0.15–0.73; range: 0–2.29). Among asymptomatic individuals, the median was 0.44 (IQR = 0.17–0.69; range: 0–1.78). Across all clusters, the median number of onward infections was similar between groups (Wilcoxon p = 0.56) (Figure 4). When restricting the analysis to incarcerated individuals only, we found no significant difference in estimated average number of secondary infections between symptomatic and asymptomatic individuals. Among asymptomatic individuals, the median number of secondary infections was 0.30 (IQR: 0.06–0.47), and among symptomatic individuals, the median was 0.21 (IQR: 0.04–0.40) (Wilcoxon p = 0.27). To illustrate transmission patterns, we visualized the inferred network for Cluster 6 (the largest genomic cluster in our dataset (n = 315) highlighting the direction and probability of transmission events by symptom status (Figure 5). Similar network visualizations for three additional large clusters are provided in the Supplementary Material (Figure S3; Figure S4 and Figure S5).

**Figure 4.**
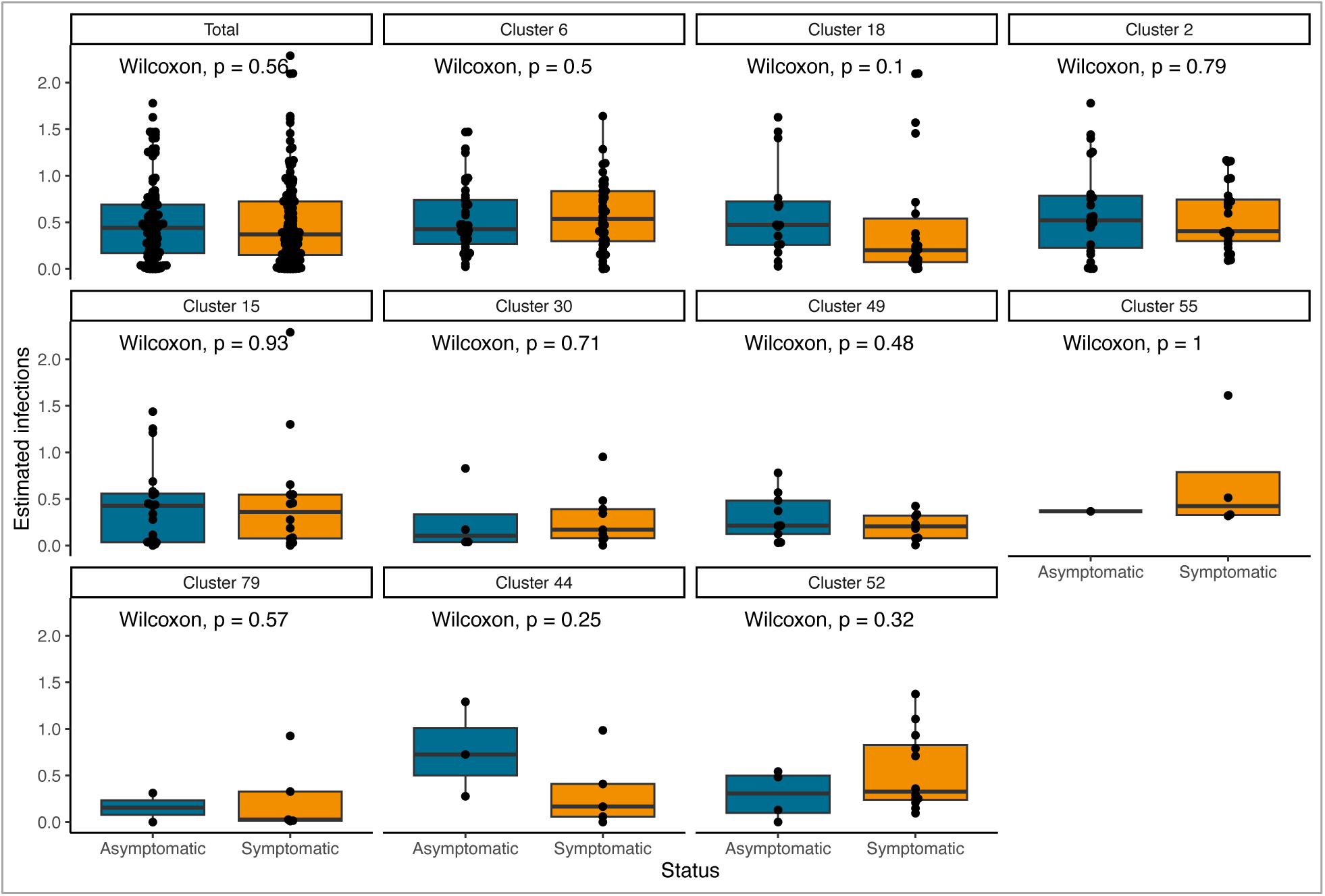
Contribution of symptomatic and asymptomatic individuals to TB transmission. Estimated number of secondary infections attributed to asymptomatic and symptomatic individuals, inferred from transmission trees using the BREATH model. Each panel represents either the total dataset or a specific transmission cluster, with boxplots comparing the number of estimated secondary cases by symptom status. The Wilcoxon rank-sum test was used to assess statistical differences between groups, and p-values are shown for each comparison.

**Figure 5.**
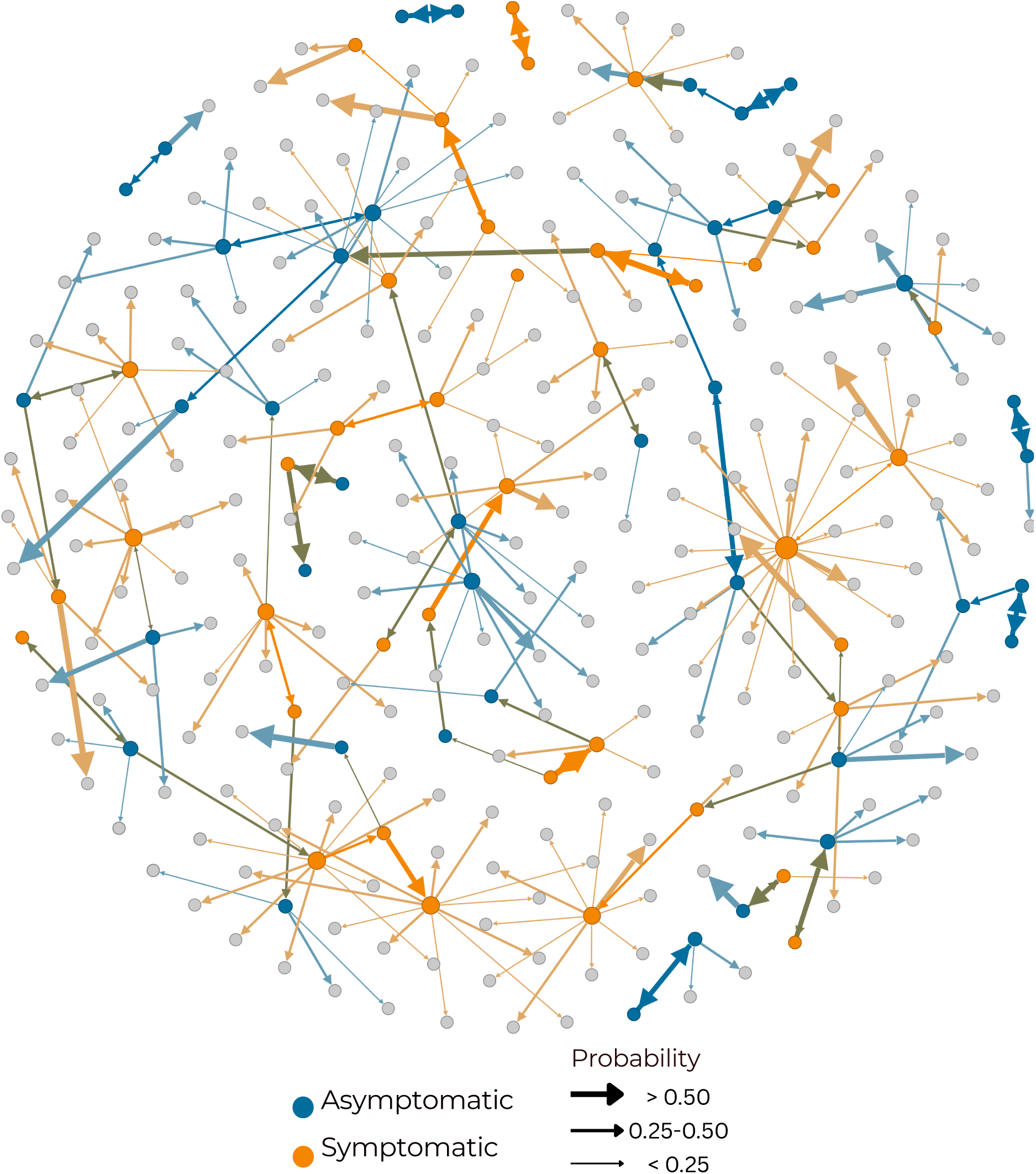
Transmission network for Cluster 6, the largest genomic cluster in the dataset (n = 315), reconstructed from posterior transmission trees inferred using the BREATH model. Each node represents a sampled individual; symptomatic individuals are shown as orange circles and asymptomatic individuals as teal circles. Individuals with unknown symptom status are represented by gray circles. Directed arrows indicate the most probable “who-infected-whom” relationships, inferred from the posterior distribution. Arrow thickness reflects the posterior probability of transmission: thick arrows represent probabilities >0.50, medium arrows 0.25– 0.50, and thin arrows <0.25.

## Discussion

This analysis integrates genomic, epidemiological, and clinical data to explore the role of asymptomatic tuberculosis in transmission, leveraging population-based surveillance including active case finding within an incarcerated populations. Using multiple analytic frameworks, including genomic clustering, phylogenetic metrics of epidemic success, and probabilistic transmission trees we observed no significant difference in measures of transmission between symptomatic and asymptomatic individuals. These results add to a growing body of evidence that individuals with asymptomatic tuberculosis contribute substantially to transmission, comparable to that of symptomatic individuals.

Multiple recent studies have demonstrated that, at a given time point, approximately half of all individuals with pulmonary tuberculosis lack symptoms (18). Modeling studies have indicated that such individuals may contribute substantially to transmission (4) and that case-finding strategies that rely on detection of symptomatic individuals may be insufficient to achieve substantive reductions in tuberculosis incidence (19). However, there is little empirical data on the relative infectiousness of individuals according to their symptoms or their overall contribution to *M. tuberculosis* transmission. Studies have found similar risk of tuberculosis infection, as measured by tuberculin skin tests or interferon gamma release assays, in close contacts of symptomatic and asymptomatic tuberculosis cases (20,21); however, such approaches have limitations in that the outcome (*M. tuberculosis* infection) reflects cumulative life exposures but was measured cross-sectionally. Further, studies indicate that much of *M. tuberculosis* transmission occurs outside of close contact networks (22). A genomic epidemiology study from the Valencia region of Spain, using a related transmission inferential approach as in our study, estimated that 5 of 14 individuals with tuberculosis likely transmitted *M. tuberculosis* before onset of symptoms. Our results build upon these earlier studies, providing additional evidence from a large genomic surveillance study with an active case finding component, which mitigates the effects of symptoms on diagnosis.

These findings highlight the importance of identifying individuals with asymptomatic TB and expanding active case-finding strategies beyond symptom-based approaches. Incorporating radiographic screening and molecular diagnostics is essential for detecting cases earlier and interrupting transmission. While this is especially critical in incarcerated populations, where transmission is amplified, it also has important implications for the broader community, where undetected asymptomatic cases may sustain TB epidemics.

Our findings underscore the urgent need to strengthen TB detection and treatment efforts in prison settings and to raise awareness about the amplified transmission risks associated with incarceration. Isolates from incarcerated individuals were more frequently found in genomic transmission clusters providing phylogenetic evidence that recent transmission is more common in prisons than in surrounding communities. Moreover, *M. tuberculosis* strains from prisons and the community were closely related, and most transmission clusters included both incarcerated and non-incarcerated individuals. Incarceration history appeared as a consistent and strong predictor of *M. tuberculosis* epidemic success. Both THD and LBI metrics demonstrated significantly higher transmission potential among currently and formerly incarcerated individuals, independent of symptom status or bacterial lineage. These results highlight the interconnection of prison and community transmission and indicate that mitigating TB transmission within prisons is a critical public health priority with broad implications. Our findings are consistent with earlier genomic studies in high-incarceration settings, which demonstrated frequent transmission between incarcerated and non-incarcerated individuals, showing that prison-based transmission is a key driver of community-level TB burden (11,23–25). Similarly, a previous study from Mato Grosso do Sul found that over 70% of community TB cases were genetically linked to clusters that included incarcerated individuals, further supporting prisons as amplification points for TB spread (26). Our findings affirm and extend this prior work by incorporating clinical and symptom status data, and by showing that the transmission potential is not limited to symptomatic individuals.

Our findings should be interpreted within the context of the limitations of the study design and available data. First, although we sequenced all available *M. tuberculosis* cultures, the final number of genomes represents only a subset of the total notified TB cases in Mato Grosso do Sul over the study period. As a result, there is underrepresentation of sequences and some circulating genotypes may not have been captured, potentially leading to an underestimation of the true extent of recent transmission. Second, symptom data were only available for a subset of individuals who were identified through active case finding, as a result, our findings reflect transmission dynamics in a high incidence setting which may limit the generalizability of comparisons between symptomatic and asymptomatic cases. Third, *M. tuberculosis* transmission rates are much higher in prisons than in the general population, and the relative contribution of symptomatic versus asymptomatic infections may differ in populations with lower transmission rates. Further, symptom status and reporting in incarcerated individuals, who are mostly young men with a high prevalence of tobacco use, are not likely representative of those in the broader population. Fourth, a key limitation of this and other studies evaluating infectiousness according to symptoms is that symptoms are evaluated at a single timepoint—at the time diagnosis— whereas evidence suggests they are highly dynamic during the course of tuberculosis. In particular, individuals with symptomatic tuberculosis at diagnosis likely had a substantial period of asymptomatic disease prior to their symptom onset (and sometimes the converse is true symptoms regressing prior to diagnosis), and it remains difficult to understand how much transmission occurred during the symptomatic versus symptomatic period. Finally, model-based transmission inference is sensitive to assumptions about transmission timing, sampling probability, and within-host diversity. While BREATH incorporates unsampled hosts and epidemiological timing, and we performed sensitivity analyses on key model parameters, the true contribution of asymptomatic individuals may still be underestimated, particularly in the presence of undiagnosed or unreported cases that might be more likely to be asymptomatic.

This study provides genomic epidemiological evidence that asymptomatic individuals play a significant role in the transmission of *M. tuberculosis*. By integrating clinical data with whole-genome sequencing and transmission modeling, we show that transmission potential is not determined by symptom presentation alone. These findings underscore the limitations of symptom-based screening and the urgent need for broader diagnostic strategies that can detect TB earlier in the disease course. Systematic screening in high-risk populations, irrespective of symptoms, will be essential for interrupting transmission and accelerating progress toward TB elimination.

## Contributors

KEdS: data curation, formal analysis, investigation, methodology, visualization, writing (original draft). JRA: conceptualization, funding acquisition, investigation, methodology, visualization, writing (review and editing), and supervision. JC: conceptualization, funding acquisition, investigation, methodology, writing (review and editing), and supervision. PCPS, DHT: data curation, methodology, and writing (review and editing). KSW, EATC, CC, TC: investigation, methodology, writing (review and editing). RDO, JVBB, MGC, CCMG, LHFD: investigation, writing (review and editing). All authors had access to the data used in the study and accept responsibility for the decision to submit for publication. KEdS and JRA have directly accessed and verified the underlying data reported in the manuscript.

## Data sharing

Raw sequence data generated in this study have been deposited in the under the study accession number PRJEB96387. The H37Rv reference genome is available on NCBI under accession number NC_000962.3.

## Ethical approval

This study was approved by the National Commission for Ethics in Research (CONEP) (3.483.377, CAAE 37237814.4.0000.5160), the Federal University of Mato Grosso do Sul Research Ethics Committee (CEP), and Stanford University. All participants provided written informed consent prior to enrollment.

## Declaration of interests

We declare no competing interests.

## Data Availability

All data produced in the present work are contained in the manuscript

## Acknowledgments

This work was supported by funding from the U.S. National Institutes of Health (R01 AI130058 and R01 AI149620) and the Brazilian National Research Council (CNPq).

## Notes

### Competing Interest Statement

The authors have declared no competing interest.

### Author Declarations

National Commission for Ethics in Research (CONEP) (3.483.377, CAAE 37237814.4.0000.5160), the Federal University of Mato Grosso do Sul Research Ethics Committee (CEP), and Stanford University gave ethical approval for this work

## References

1. World Health Organization. Global Tuberculosis Report 2024. 1st ed. World Health Organization; 2024.

2. Emery JC, Dodd PJ, Banu S, Frascella B, Garden FL, Horton KC, et al. Estimating the contribution of subclinical tuberculosis disease to transmission: An individual patient data analysis from prevalence surveys. eLife. 2023 Dec 18;12:e82469.

3. Bezerra W da SP, Jung E, Croda MG, Oliveira RD de, Santos A da S, Tsuha DH, et al. The role of tuberculosis symptoms in transmission risk to cell contacts in prisons [Internet]. medRxiv; 2025 [cited 2025 June 5]. p. 2025.05.27.25328413. Available from: https://www.medrxiv.org/content/10.1101/2025.05.27.25328413v1

4. Ryckman TS, Dowdy DW, Kendall EA. Infectious and clinical tuberculosis trajectories: Bayesian modeling with case finding implications. Proc Natl Acad Sci U S A. 2022 Dec 27;119(52):e2211045119.

5. Phelan JE, O’Sullivan DM, Machado D, Ramos J, Oppong YEA, Campino S, et al. Integrating informatics tools and portable sequencing technology for rapid detection of resistance to anti-tuberculous drugs. Genome Med. 2019 June 24;11(1):41.

6. Page AJ, Taylor B, Delaney AJ, Soares J, Seemann T, Keane JA, et al. SNP-sites: rapid efficient extraction of SNPs from multi-FASTA alignments. Microb Genomics. 2016;2(4):e000056.

7. Kalyaanamoorthy S, Minh BQ, Wong TKF, von Haeseler A, Jermiin LS. ModelFinder: fast model selection for accurate phylogenetic estimates. Nat Methods. 2017 June;14(6):587–9.

8. Paradis E, Schliep K. ape 5.0: an environment for modern phylogenetics and evolutionary analyses in R. Bioinformatics. 2019 Feb 1;35(3):526–8.

9. Guerra-Assunção JA, Crampin AC, Houben RMGJ, Mzembe T, Mallard K, Coll F, et al. Large-scale whole genome sequencing of M. tuberculosis provides insights into transmission in a high prevalence area. eLife. 2015 Mar 3;4:e05166.

10. Rasigade JP, Barbier M, Dumitrescu O, Pichat C, Carret G, Ronnaux-Baron AS, et al. Strain-specific estimation of epidemic success provides insights into the transmission dynamics of tuberculosis. Sci Rep. 2017 Mar 28;7:45326.

11. Sanabria GE, Sequera G, Aguirre S, Méndez J, dos Santos PCP, Gustafson NW, et al. Phylogeography and transmission of Mycobacterium tuberculosis spanning prisons and surrounding communities in Paraguay. Nat Commun. 2023 Jan 19;14(1):303.

12. Neher RA, Russell CA, Shraiman BI. Predicting evolution from the shape of genealogical trees. McVean G, editor. eLife. 2014 Nov 11;3:e03568.

13. Fischer M, Herbst L, Kersting S, Kühn AL, Wicke K. Tree Balance Indices: A Comprehensive Survey [Internet]. Cham: Springer International Publishing; 2023 [cited 2025 Apr 9]. Available from: https://link.springer.com/10.1007/978-3-031-39800-1

14. Andrews JR, Vaidya K, Bern C, Tamrakar D, Wen S, Madhup S, et al. High Rates of Enteric Fever Diagnosis and Lower Burden of Culture-Confirmed Disease in Peri-urban and Rural Nepal. J Infect Dis. 2018 Nov 10;218(suppl_4):S214–21.

15. Bouckaert R, Heled J, Kühnert D, Vaughan T, Wu CH, Xie D, et al. BEAST 2: A Software Platform for Bayesian Evolutionary Analysis. PLOS Comput Biol. 2014 Apr 10;10(4):e1003537.

16. Colijn C, Hall M, Bouckaert R. Taking a BREATH (Bayesian Reconstruction and Evolutionary Analysis of Transmission Histories) to simultaneously infer phylogenetic and transmission trees for partially sampled outbreaks [Internet]. bioRxiv; 2024 [cited 2025 Apr 9]. p. 2024.07.11.603095. Available from: https://www.biorxiv.org/content/10.1101/2024.07.11.603095v1

17. Menardo F, Duchêne S, Brites D, Gagneux S. The molecular clock of Mycobacterium tuberculosis. PLoS Pathog. 2019 Sept;15(9):e1008067.

18. Stuck L, Klinkenberg E, Abdelgadir Ali N, Basheir Abukaraig EA, Adusi-Poku Y, Alebachew Wagaw Z, et al. Prevalence of subclinical pulmonary tuberculosis in adults in community settings: an individual participant data meta-analysis. Lancet Infect Dis. 2024 July;24(7):726–36.

19. Dowdy DW, Basu S, Andrews JR. Is passive diagnosis enough? The impact of subclinical disease on diagnostic strategies for tuberculosis. Am J Respir Crit Care Med. 2013 Mar 1;187(5):543–51.

20. Emery JC, Dodd PJ, Banu S, Frascella B, Garden FL, Horton KC, et al. Estimating the contribution of subclinical tuberculosis disease to transmission: An individual patient data analysis from prevalence surveys. eLife. 2023 Dec 18;12:e82469.

21. Bezerra W da SP, Jung E, Croda MG, Oliveira RD de, Santos A da S, Tsuha DH, et al. The role of tuberculosis symptoms in transmission risk to cell contacts in prisons [Internet]. medRxiv; 2025 [cited 2025 June 18]. p. 2025.05.27.25328413. Available from: https://www.medrxiv.org/content/10.1101/2025.05.27.25328413v1

22. Martinez L, Lo NC, Cords O, Hill PC, Khan P, Hatherill M, et al. Paediatric tuberculosis transmission outside the household: challenging historical paradigms to inform future public health strategies. Lancet Respir Med. 2019 June;7(6):544–52.

23. Roycroft E, Fitzgibbon MM, Kelly DM, Scully M, McLaughlin AM, Flanagan PR, et al. The largest prison outbreak of TB in Western Europe investigated using whole-genome sequencing. Int J Tuberc Lung Dis Off J Int Union Tuberc Lung Dis. 2021 June 1;25(6):491– 7.

24. Miyahara R, Piboonsiri P, Chiyasirinroje B, Imsanguan W, Nedsuwan S, Yanai H, et al. Risk for Prison-to-Community Tuberculosis Transmission, Thailand, 2017–2020 - Volume 29, Number 3—March 2023 - Emerging Infectious Diseases journal - CDC. [cited 2025 May 9]; Available from: https://www.nc.cdc.gov/eid/article/29/3/22-1023_article

25. Utpatel C, Zavaleta M, Rojas-Bolivar D, Mühlbach A, Picoy J, Portugal W, et al. Prison as a driver of recent transmissions of multidrug-resistant tuberculosis in Callao, Peru: a cross-sectional study. Lancet Reg Health – Am [Internet]. 2024 Mar 1 [cited 2025 May 9];31. Available from: https://www.thelancet.com/journals/lanam/article/PIIS2667-193X%2824%2900001-2/fulltext?utm_source=chatgpt.com

26. Walter KS, Santos PCP dos, Gonçalves TO, Silva BO da, Santos A da S, Leite A de C, et al. The role of prisons in disseminating tuberculosis in Brazil: A genomic epidemiology study. Lancet Reg Health – Am [Internet]. 2022 May 1 [cited 2025 Mar 17];9. Available from: https://www.thelancet.com/journals/lanam/article/PIIS2667-193X(22)00003-5/fulltext

