## Supplemental material for "Comparison of phylogenetic metrics of transmission in symptomatic and asymptomatic tuberculosis"

The supplementary material includes supplementary text (S1 Text, Supplementary Methods), Figures S1-S5, and Supplementary References.

| | |
|---|---|
| S1 Text. Supplementary Methods | 2 |
| Figure. S1 | 5 |
| Figure. S2 | 6 |
| Figure. S3 | 7 |
| Figure. S4 | 8 |
| Figure. S5 | 9 |
| Supplementary References | 10 |

### S1 Text. Supplementary Methods

#### Study population

All participants provided written informed consent and completed a structured questionnaire covering prior tuberculosis diagnosis, treatment history and outcomes, potential contact with individuals diagnosed with pulmonary tuberculosis, and incarceration history. Additional sociodemographic and clinical data were obtained from the notifiable disease registry (SINAN). Incarceration history was obtained from the state incarceration database (SIGO), and individuals classified as currently incarcerated, previously incarcerated, or no history of incarceration, according to their status at the time of their tuberculosis diagnosis.

#### Laboratory diagnosis and culture

Sputum samples were cultured using the Ogawa-Kudoh method (1). Cultures were incubated at 37 °C and monitored twice weekly for up to 60 days. DNA was extracted from positive *Mycobacterium tuberculosis* cultures using the cetyltrimethylammonium bromide (CTAB) method (2).

#### Whole genome sequencing and bioinformatics analyses

Low-quality bases (Phred score <20) were trimmed, and adapters were removed using Trim Galore v0.6.5 (stringency=3). Additional filtering was performed with CutAdapt v4.2 (–nextseq-trim=20, –minimum-length=20, –pair-filter=any) (3). To minimize contamination, reads were taxonomically classified using Kraken2 (4), and only those assigned to the *Mycobacterium* genus and specifically to *M. tuberculosis* were retained. Reads were aligned to the *M. tuberculosis*

H37Rv reference genome (NCBI Accession: NC_000962.3) using BWA v0.7.15 (5), and duplicate reads were removed using Sambamba (6). Variant calling was performed using GATK v4.1 with HaplotypeCaller (sample ploidy=1) and GenotypeGVCFs (7). We retained variants with a minimum depth of 10× and a quality score of at least 40, including non-variant sites in the final VCF files. Consensus sequences were constructed using bcftools consensus (8), excluding indels. SNPs located in repetitive regions (e.g., PPE and PE-PGRS genes, phages, insertion sequences, and repeats >50 bp) were excluded from downstream analyses (9).

**Phylogenetic reconstruction**

To construct the phylogeny, we identified the best-fit substitution model using ModelFinder implemented in IQ-TREE v2.2.0, evaluating all models that included ascertainment bias correction appropriate for SNP-only alignments (10). Based on the Bayesian Information Criterion (BIC), the best-fit model was TVM+F+ASC+R7. Phylogenetic trees were visualized using iTOL (Interactive Tree of Life) (11).

**Transmission inference**

To explore how different assumptions about the timing of transmission events affect the results, we ran the model under two prior settings for the transmission rate. These priors reflect assumptions about the relative speed of transmission compared to case detection. In the "long" scenario, we set the mean transmission interval to 2.0 years, representing faster transmission relative to sampling. In the "medium" scenario, the mean transmission interval was 1.5 years, representing a shorter time between infection events and case detection. We corrected for ascertainment bias by specifying the number of invariant sites in the BEAST XML configuration, using the R package beautier (https://docs.ropensci.org/beautier/). Markov chain Monte Carlo (MCMC) chains were run for 200 million iterations, or longer if needed for convergence, discarding the first 10% of samples as burn-in. Convergence was assessed using Tracer v1.7.1, confirming that all model parameters had effective sample sizes (ESS) > 200. TreeAnnotator v2.7.6 was used to summarize the posterior tree distributions (12).

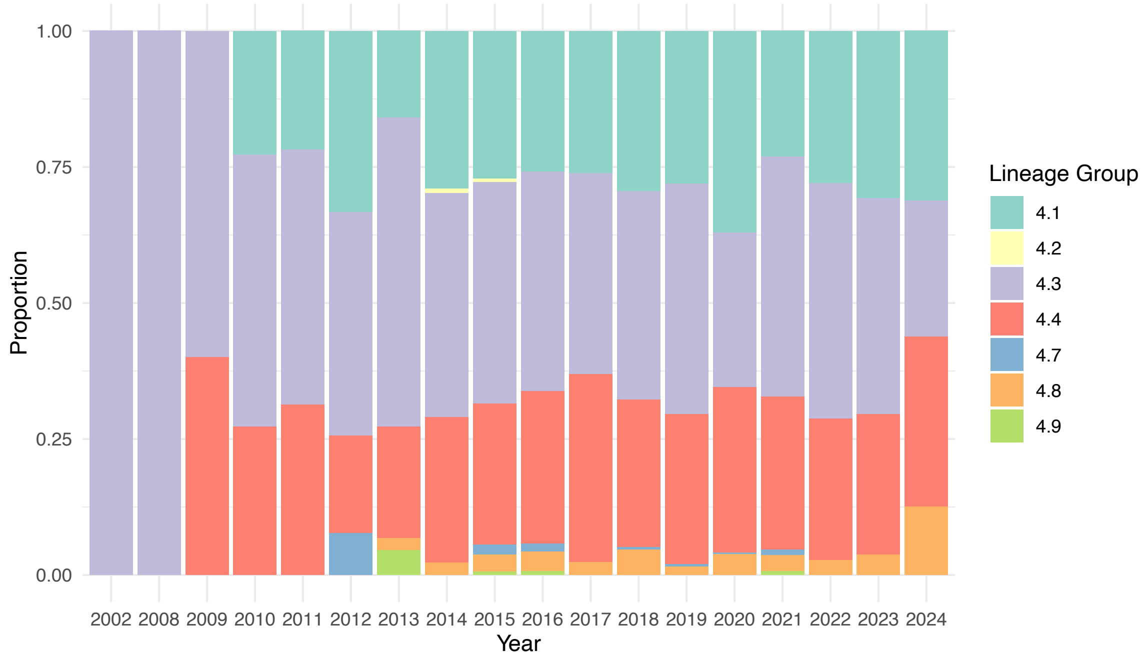


**Figure S1. Genomic surveillance of *M. tuberculosis* in Mato Grosso do Sul, Brazil, 2002-2024**. Proportion of genomes of lineage 4 sub-lineages collected during the study period.

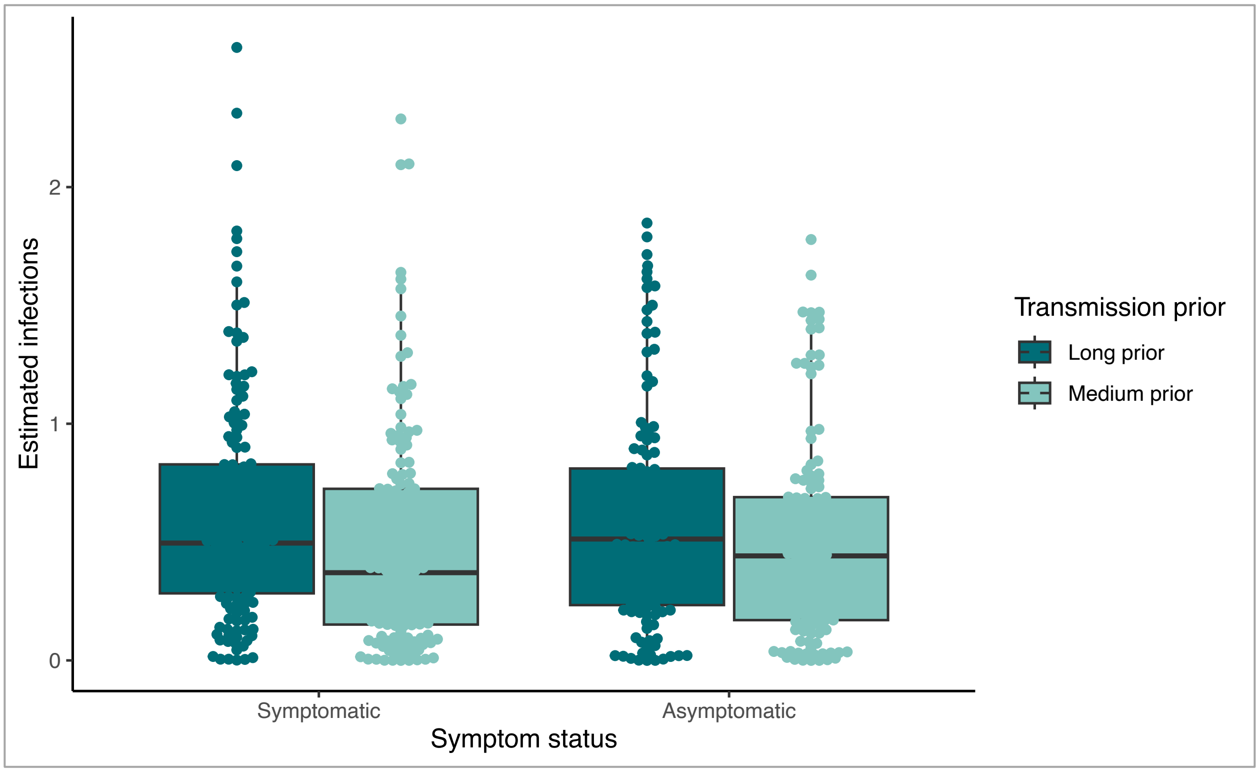


**Figure S2. Estimated number of secondary infections attributed to symptomatic and asymptomatic individuals under two transmission prior scenarios.** Boxplots show the distribution of inferred secondary infections per individual based on transmission trees reconstructed using the BREATH model. Estimates are shown separately for the "long" prior (mean transmission interval = 2.0 years) and the "medium" prior (mean = 1.5 years). Symptomatic and asymptomatic individuals are represented on the x-axis. Slight differences between priors are visible, but overall patterns remain consistent across symptom status groups.

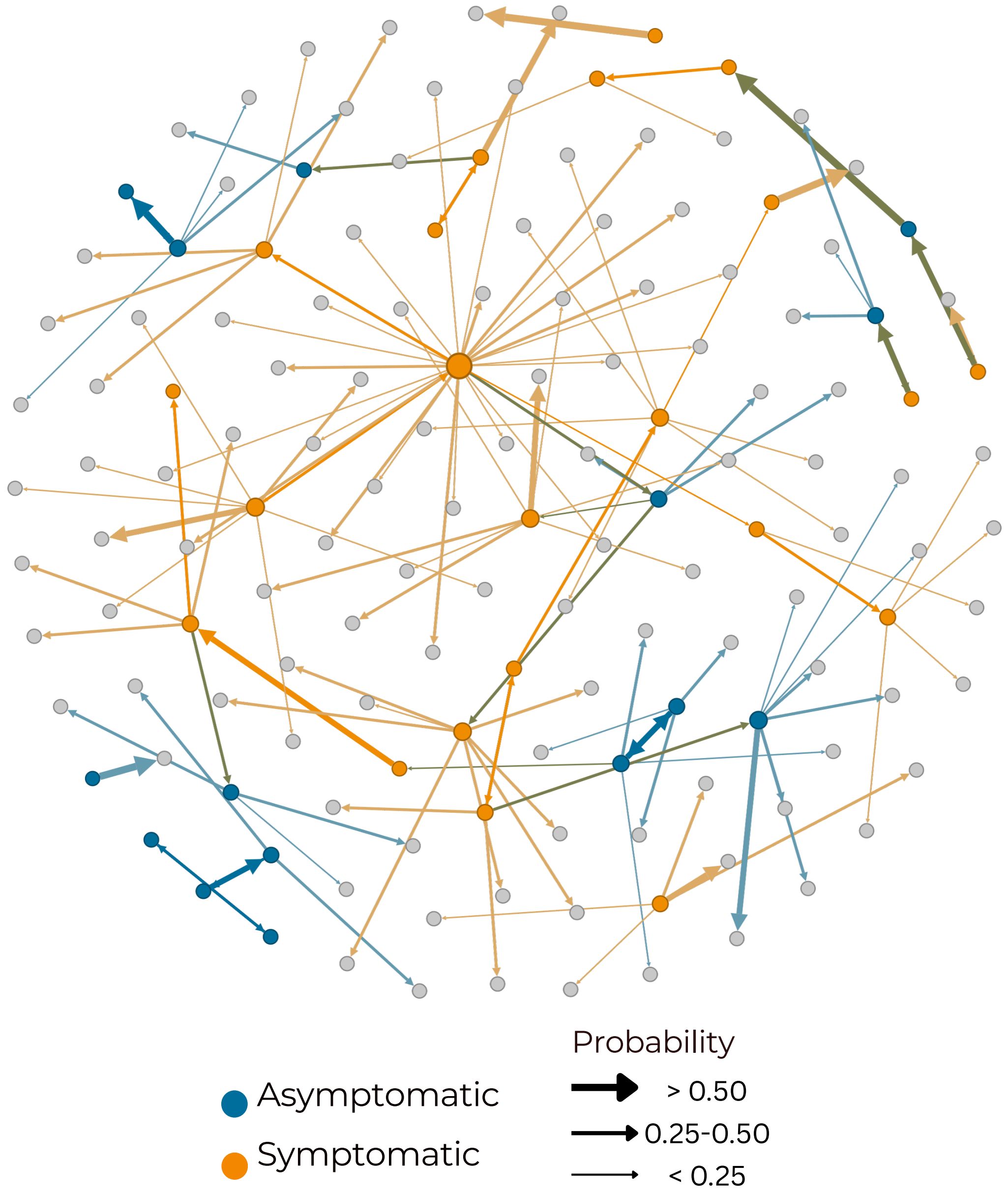


**Figure S3. Transmission network for Cluster 18 (n = 167), reconstructed from posterior transmission trees inferred using the BREATH model.** Each node represents a sampled individual; symptomatic individuals are shown as orange circles and asymptomatic individuals as teal circles. Directed arrows indicate the most probable "who-infected-whom" relationships, inferred from the posterior distribution. Arrow thickness reflects the posterior probability of transmission: thick arrows represent probabilities >0.50, medium arrows 0.25–0.50, and thin arrows <0.25.

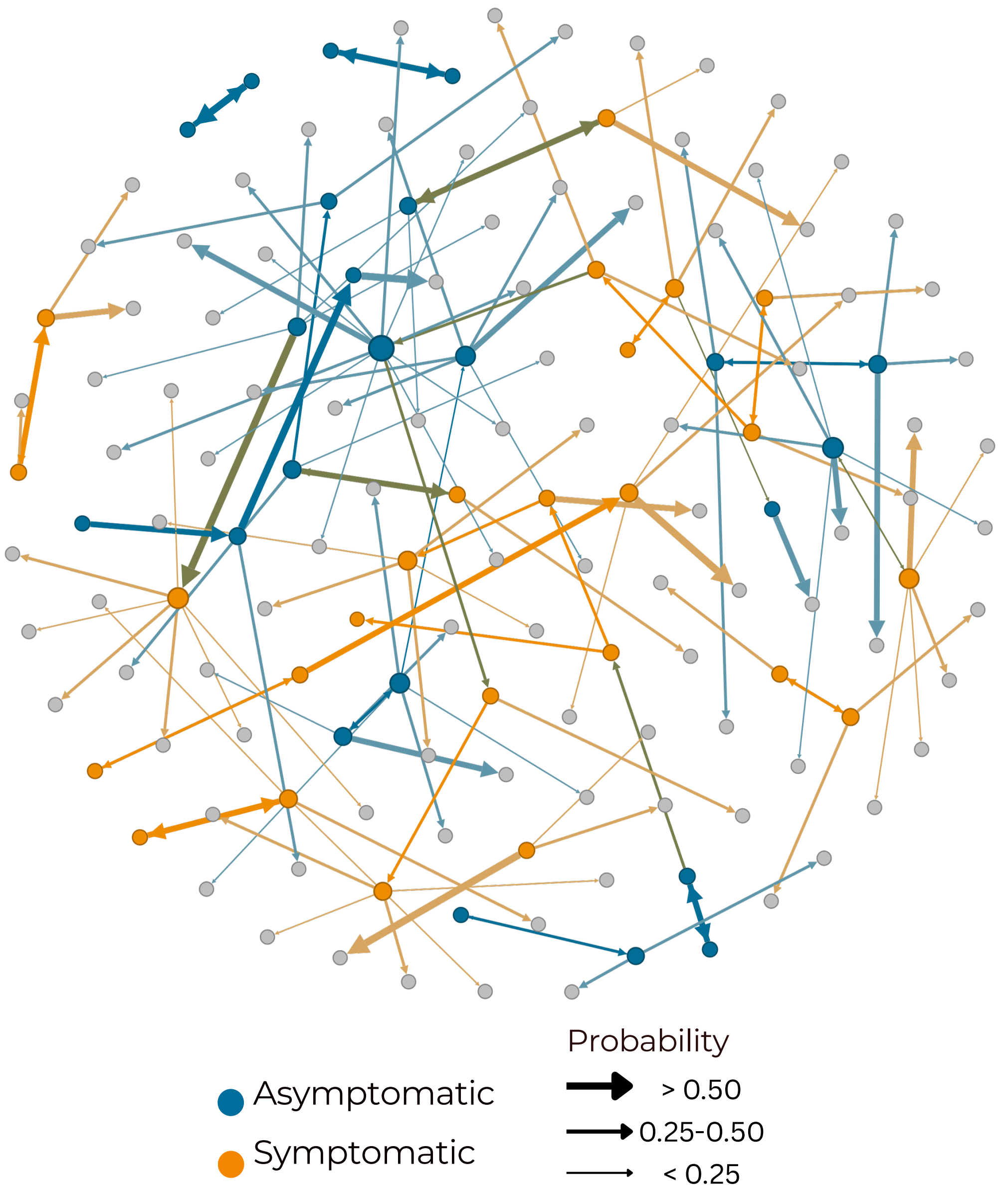


**Figure S4. Transmission network for Cluster 2 (n = 152), reconstructed from posterior transmission trees inferred using the BREATH model.** Each node represents a sampled individual; symptomatic individuals are shown as orange circles and asymptomatic individuals as teal circles. Directed arrows indicate the most probable "who-infected-whom" relationships, inferred from the posterior distribution. Arrow thickness reflects the posterior probability of transmission: thick arrows represent probabilities >0.50, medium arrows 0.25–0.50, and thin arrows <0.25.

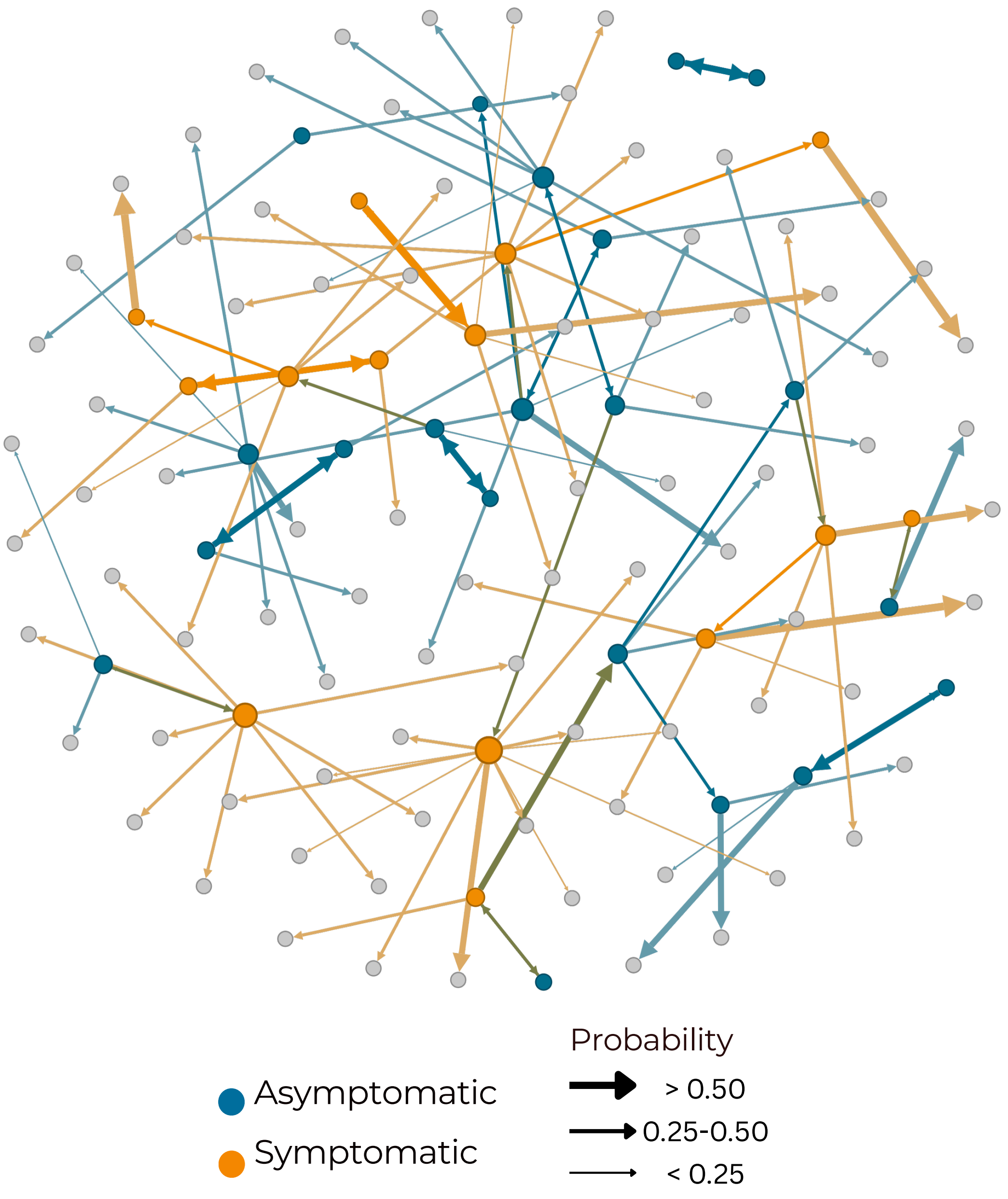


**Figure S5. Transmission network for Cluster 15 (n = 128), reconstructed from posterior transmission trees inferred using the BREATH model.** Each node represents a sampled individual; symptomatic individuals are shown as orange circles and asymptomatic individuals as teal circles. Directed arrows indicate the most probable "who-infected-whom" relationships, inferred from the posterior distribution. Arrow thickness reflects the posterior probability of transmission: thick arrows represent probabilities >0.50, medium arrows 0.25–0.50, and thin arrows <0.25.

**Supplementary References**


1. Palaci M, Peres RL, Maia R, Cunha EAT, Ribeiro MO, Lecco R, et al. Contribution of the Ogawa-Kudoh swab culture method to the diagnosis of pulmonary tuberculosis in Brazil. Int J Tuberc Lung Dis. 2013 June 1;17(6):782–6.

2. Schiebelhut LM, Abboud SS, Gómez Daglio LE, Swift HF, Dawson MN. A comparison of DNA extraction methods for high-throughput DNA analyses. Mol Ecol Resour. 2017;17(4):721–9.

3. Martin M. Cutadapt removes adapter sequences from high-throughput sequencing reads. EMBnet.journal. 2011 May 2;17(1):10–2.

4. Wood DE, Lu J, Langmead B. Improved metagenomic analysis with Kraken 2. Genome Biol. 2019 Nov 28;20(1):257.

5. Li H, Durbin R. Fast and accurate short read alignment with Burrows–Wheeler transform. Bioinformatics. 2009 July 15;25(14):1754–60.

6. Tarasov A, Vilella AJ, Cuppen E, Nijman IJ, Prins P. Sambamba: fast processing of NGS alignment formats. Bioinformatics. 2015 June 15;31(12):2032–4.

7. Poplin R, Ruano-Rubio V, DePristo M, Fennell T, Carneiro M, Van der Auwera G, et al. Scaling accurate genetic variant discovery to tens of thousands of sample [Internet]. 2017 [cited 2025 Apr 9]. Available from: https://doi.org/10.1101/201178

8. Danecek P, Bonfield JK, Liddle J, Marshall J, Ohan V, Pollard MO, et al. Twelve years of SAMtools and BCFtools. GigaScience. 2021 Feb 1;10(2):giab008.

9. Brites D, Loiseau C, Menardo F, Borrell S, Boniotti MB, Warren R, et al. A New Phylogenetic Framework for the Animal-Adapted Mycobacterium tuberculosis Complex. Front Microbiol [Internet]. 2018 Nov 27 [cited 2025 Apr 9];9. Available from: https://www.frontiersin.org/journals/microbiology/articles/10.3389/fmicb.2018.02820/full

10. Minh BQ, Schmidt HA, Chernomor O, Schrempf D, Woodhams MD, von Haeseler A, et al. IQ-TREE 2: New Models and Efficient Methods for Phylogenetic Inference in the Genomic Era. Mol Biol Evol. 2020 May 1;37(5):1530–4.

11. Letunic I, Bork P. Interactive Tree of Life (iTOL) v6: recent updates to the phylogenetic tree display and annotation tool. Nucleic Acids Res. 2024 July 5;52(W1):W78–82.

12. Bilderbeek RJC, Etienne RS. babette: BEAUti 2, BEAST2 and Tracer for R. Methods Ecol Evol. 2018;9(9):2034–40.
